# GEM-Finder: dissecting GWAS variants via long-range interacting cis-regulatory elements with differentiation-specific genes

**DOI:** 10.1101/2025.03.02.25323165

**Authors:** Gyeongsik Park, Andrew J. Lee, Inkyung Jung

## Abstract

Interpreting the functional significance of non-coding GWAS variants remains challenging. While co-localizing variants with cell-type specific cis-regulatory elements (CREs) has improved our understanding, many variants remain unassociated. In this study, we propose GEM-Finder (Genomic Element Mapping for Fine Discovery of Promoter-Linked Variants), a novel analytical framework that integrates transcriptomic, epigenomic (H3K27ac ChIP-seq), and chromatin interaction data. GEM-Finder utilizes long-range chromatin interactions to identify CREs that connect differentially expressed genes of specific cell types. When we apply GEM-Finder to endothelial differentiation, unlike conventional methods primarily focused on cell-type specific CREs, GEM-Finder identifies 7.6 times more disease/trait associations. Specifically, by integrating transcriptome, epigenome (particularly H3K27ac ChIP-seq), and long-range chromatin interactions during endothelial differentiation, we identified CREs linked to differentiation-specific genes. Our enrichment analyses revealed both shared and unique associations for 53 human diseases/traits. Notably, the majority of these (68%) exhibited unique associations in a differentiation-specific manner. Hematological traits and neuropsychiatric disorders were primarily linked to the final stage of endothelial differentiation, while several complex diseases, such as colorectal cancer (CRC), were unexpectedly associated with the late stage. Our findings underscore the importance of leveraging long-range chromatin interactions to accurately identify disease-associated CREs in the functional characterization of non-coding GWAS variants.

## Introduction

The advent of genome-wide association studies (GWAS) has transformed our understanding of the genetic foundations of complex human diseases/traits [1]. While these studies have identified numerous non-coding genetic variants associated with various conditions, interpreting the function of these variants remains a significant challenge, as the majority of GWAS variants reside in non-coding genomic regions [2]. To address the gap between non-coding GWAS variants and their related molecular phenotypes, cis-regulatory elements (CREs), regulatory DNA sequences that can influence gene expression over long distances, have emerged as critical mediators of non-coding variant effects [3]. CREs tightly control spatiotemporal gene regulation by modulating the transcriptional activities of target promoters through physical interactions [4-6]. Therefore, disease-risk genetic variants found within cell-type specific CREs potentially alter specific transcription factor (TF) binding, consequently disrupting cell-type specific gene regulatory programs [7-8].

However, characterizing the regulatory impact of GWAS variants remains challenging, especially since many GWAS variants are not located within cell-type specific CREs, complicating the functional interpretation of these non-coding variants. To address this, we leveraged long-range chromatin interactions to identify CREs engaged with cell-type specific genes, enabling us to rescue additional CREs that harbor GWAS SNPs beyond just cell-type specific CREs solely with ChIP-seq. While there is a strong association between cell-type specific CREs and gene expression [9][10], a substantial number of shared CREs between cell types are also linked to cell-type specific gene regulation, suggesting that considering only stage-specific CREs may overlook many important genetic variants. In this regard, we hypothesized that CREs physically connecting to specific promoters via long-range chromatin interactions may provide more comprehensive and direct association with disease-specific target genes than those presenting solely cell-type specific activities. To test our hypothesis, we proposed a new analytical framework with variant multi-omics data called <u>G</u>enomic <u>E</u>lement <u>M</u>apping for <u>Fin</u>e Discov<u>er</u>y of Promoter-Linked Variants (GEM-Finder). GEM-Finder is a conceptually innovative computational approach for prioritizing cell types or differentiation stages associated with specific human diseases and traits. Unlike conventional approaches that primarily based on the colocalization of GWAS variants within cell-type-specific genes or CREs such as SNPsea [11], cell-type-specific DHS regions [12], LDSC [13], and LDSC-SEG [14], GEM-Finder emphasizes CREs linked to stage-specific DEGs based on the assumption that many CREs without stage specificity can still contribute to stage-specific DEG expression.

We employed GEM-Finder to explore complex human diseases/traits associated with endothelial cells (ECs). ECs, which line the entire circulatory system, are essential for maintaining bodily homeostasis, including blood pressure regulation, immune cell trafficking, oxygen and nutrient transport, stem cell maintenance, and the creation of tissue-specific microenvironments [15-18]. Endothelial dysfunction can impair vasodilation and promote a pro-inflammatory, pro-thrombotic state, leading to various pathological outcomes such as cardiovascular diseases, diabetes, and chronic renal failure [19-21]. However, a systematic examination of GWAS variants about EC differentiation has yet to be conducted. By utilizing multi-omics approaches and incorporating transcriptome, epigenome, and 3D genome data during EC differentiation, GEM-Finder successfully identified unique associations with various complex human diseases/traits at each differentiation stage. Notably, we observed unexpected enrichment of GWAS variants related to colorectal cancer within the gene regulatory program specific to late-stage ECs. In conclusion, GEM-Finder underscores the value of cell-type/differentiation-specific long-range chromatin interactions with CREs in the functional characterization of GWAS variants.

## Materials and Methods

### Identification of stage-specific CREs and stage-specific DEGs in hESC-EC differentiation

We employed the quasi-likelihood F-test in EdgeR [22] to identify stage-specific CREs associated with hESC-EC differentiation based on H3K27ac ChIP-seq data. First, stage-specific CRE candidates for hESC, mesoderm, and EC were identified by iteratively contrasting each distinct cell type against the others, using a significance threshold of Benjamini-Hochberg (BH) corrected P < 0.05. Next, samples from individual differentiation stages (early, mid, and late) were iteratively compared to mesoderm to identify differentiation-dependent CREs, applying the same BH-corrected significance threshold (P < 0.05). To determine the final set of stage-specific CREs, each candidate CRE was uniquely assigned to the most significant stage based on its BH-corrected p-value.

To identify stage-specific DEGs associated with hESC-EC differentiation, we applied the quasi-likelihood F-test in the same manner as for defining stage-specific CREs but without the unique assignment step. After BH correction, we filtered each stage’s candidate genes based on an absolute expression fold change > 1.

### *In situ* Hi-C data processing

The sequencing output from Hi-C libraries was mapped to the reference genome (hg19) using BWA-mem [23] (‘–M’ option). In-house scripts were used to remove low-quality reads (MAPQ < 10), the reads that span ligation sites, chimeric reads, and self-interacting reads (two fragments located within 5kb). The chimeric reads were removed since they are by-products of proximity ligation during Hi-C experiments and cannot be properly processed by paired-end BWA-mem command. The read-pairs were merged as paired-end aligned BAM files, and PCR duplicates were removed with Picard (v2.6.0). To evaluate contact-matrix similarity across Hi-C samples, stratum-adjusted correlation coefficients S The Hi-C contact matrices were generated in .hic file format by using juicer [24] and Knight-Ruiz normalization. The topologically associating domains (TADs) were systematically identified based on normalized 40kb resolution Hi-C contact maps by processing the directionality index (DI) scores (500kb window) with Domain Caller [25]. Then, the genome was partitioned into TADs, boundaries, and unstructured regions. The unstructured regions were defined when the boundary was larger than 400 kb.

### Compartment analysis

Identification of compartment A/B was performed similarly to an earlier work [26].

### GEM-Finder framework

The key concept of GEM-Finder is the utilization of DEG-linked CRE. To address our goal, processed transcriptome, epigenome (H3K27ac), and 3D genome data are integrated through the following four steps: 1) Annotate stage-specific DEGs for each cell-type or stages using quasi-likelihood F-test with RNA-seq gene expression. 2) Identify genomic regions interacting with stage-specific DEG promoters based on long-range chromatin interactions. 3) Collect CREs defined in each cell type or stage that co-localize within stage-specific DEG-linked genomic regions. 4) Conduct GWAS variant enrichment analysis to prioritize cell type or differentiation stages associated with specific human diseases/traits. The overview of GEM-Finder workflow is in Figure 3A. Required multi-omics data were preprocessed using the aforementioned methods, and a permutation test was used for variant enrichment analysis after the stage-specific DEG-linked CREs identification.

### GWAS heritability analysis

GWAS-SNPs were obtained from the GWAS catalog database (All associations v1.0, downloaded on Aug. 2024) [27]. The genomic coordinates for GWAS-SNPs were lifted to hg19 using the UCSC liftover tool and selected 541 human traits and diseases whose number of significant tag SNPs (*P* < 5 × 10^−8^) exceeds 100. The 360,079 representative genetic variants for the 541 GWAS traits were expanded using the linkage disequilibrium (LD) structure. LD scores were calculated using PLINK for five different populations, including African (AFR), American (AMR), East Asian (EAS), European (EUR), and South Asian (SAS), from 1000 genome phase 3 data. For each tag SNP, we identified associated SNPs that share a tight LD score (r^2^ > 0.8) in at least two ethnic groups. To evaluate the statistical enrichment of GWAS-SNPs in target CREs, we considered the number of CREs in the input CREs group that are associated with linkage disequilibrium expanded GWAS-SNPs and assessed the relative enrichment compared to the random expectation individually for selected human diseases/traits. For the random expectation, we generated a random set of genome coordinates that matched the input CREs in number, size, and chromosomes. The enrichment was measured based on iterative trials (n = 1 × 10^5^) of random simulations, and the statistical significance was calculated in the form of an empirical *P*-value. This identified 53 human traits and diseases based on a significance threshold of BH corrected *P* < 0.005 and fold change over expectation > 2.

## RESULTS

### 3D genome reorganization coordinated with stage-specific gene expression and CRE activities during hESC-EC differentiation

To investigate whether 3D genome reorganization is involved in the dynamic gene regulation during endothelial differentiation from human embryonic stem cells (hESCs), we obtained multi-omics data, including transcriptome (RNA-seq), histone H3 lysine 27 acetylation (H3K27ac ChIP-seq), and 3D genome organization (Hi-C) from the GEO database (GSE281670) (**Fig. 1A and B**). In brief, the multi-omics data were generated by harvesting cells from hESCs, transitioning through the mesoderm stage by treating with GSK3β inhibitors for 72 hours, and then inducing EC differentiation with VEGF-A and forskolin for 48 hours [28]. Differentiating cells were collected at hESCs, mesoderm (ME), early (4hr), mid (12hr), late (24hr), and full (48hr) stages during EC specification with non-VEGF-A treated controls in late (24hr) and full (48hr) stages, and subjected to produce multi-omics results (**Fig. 1A**). This comprehensive dataset initially enabled us to identify a total of 136,967 CREs with ChIP-seq data, by merging H3K27ac peaks from the individual samples. For each stage, we identified 22,180 hESC-specific, 19,063 mesoderm-specific, and 42,309 EC differentiation-specific CREs (6,493 for early, 6,920 for mid, 13,452 for late, and 15,444 for full differentiation stages).

**Fig. 1.**
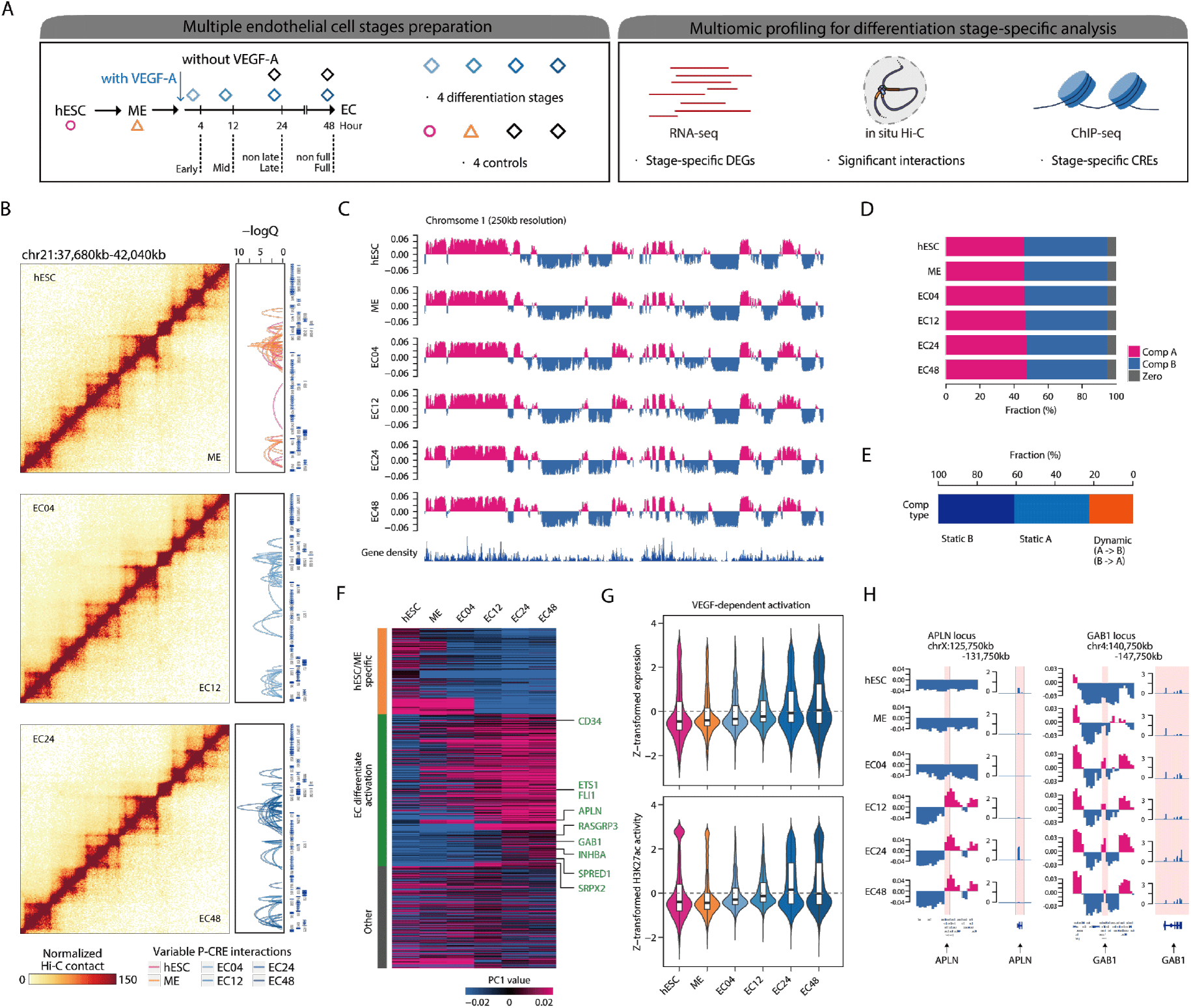
Dynamic compartment A/B patterns during EC-differentiation. **(A)** A schematic overview of EC-differentiation stage specific multiomic profiling. (**B**) Showing are Hi-C contact matrix and significant long-range interactions during EC-differentiation at x and 20kb resolution, respectively. **(C)** Shown are compartment A/B dynamics on chromosome 1 during EC-differentiation. (**D**) Stacked barplots showing the fraction of compartment A and B for each stage. (**E**) A stacked barplot showing the fraction of genomic regions subjecting to stable compartment A and B, and switching from A (or B) to B (or A). **(F)** K-mean clustering reveals hESC/ME specific and VEGF-A dependent compartment A regions. Representative genes resided in each cluster are shown together. (**G**) Violin plots showing gene expression patterns (top) and H2K27ac signals (bottom) along the EC-differentiation at VEGF-A dependent compartment A regions. **(H)** Genome browser snapshots for *APLN* and *GAB1* loci for compartment A/B patterns during EC-differentiation along with gene expression levels (TPM) at matched stage.

Next, we examined the dynamic reorganization of the 3D genome structure during EC-differentiation (**Fig. 1B**). The analysis of compartment A/B patterns, an indicator of macro-scale 3D genome organization, illustrates substantial compartment transitions in both sides during EC differentiation **(Fig.1C)**. Although overall relative ratio of compartment A and B is similar across each stage **(Fig. 1D)**, tracing each region revealed that approximately 20% of the genome switches its compartment type **(Fig. 1E)**. K-means clustering of genomic regions undergoing compartment switching revealed VEGF-A-dependent active compartment A regions, which progressively acquired during EC specification (**Fig. 1F**). Notably, these regions corresponded with upregulated gene expression and enhanced CRE activities (**Fig. 1G**). EC-progenitor (late or full stages) specific genes, such as *APLN* and *GAB1*, exhibited concordant changes between compartment A/B and gene expression, highlighting the functional role of the 3D genome organization in shaping the stage-specific transcriptome and epigenome relevant to EC specification (**Fig. 1H)**.

### Stage-specific long-range chromatin interactions involve stage-specific CREs

Since long-range chromatin interaction is a key mechanism of gene regulation, we further explored the relationship between dynamic long-range chromatin interactions and EC differentiation-specific gene expression. Using Fit-HiC [29], we identified significant long-range chromatin interactions at a 0.01 FDR cutoff for each stage, with a resolution of 20kb (**Fig. 2A**). To account for variability in the number of identified interactions across differentiation stages due to the variable Hi-C depth, we standardized the total number of significant interactions based on the sample with the lowest number of significant long-range interactions, ensuring a more reasonable approach to handling these unsaturated datasets. A total of 798,266 unique significant interactions were identified, with 74% being variable across multiple stages (**Fig. 2B**).

**Fig. 2.**
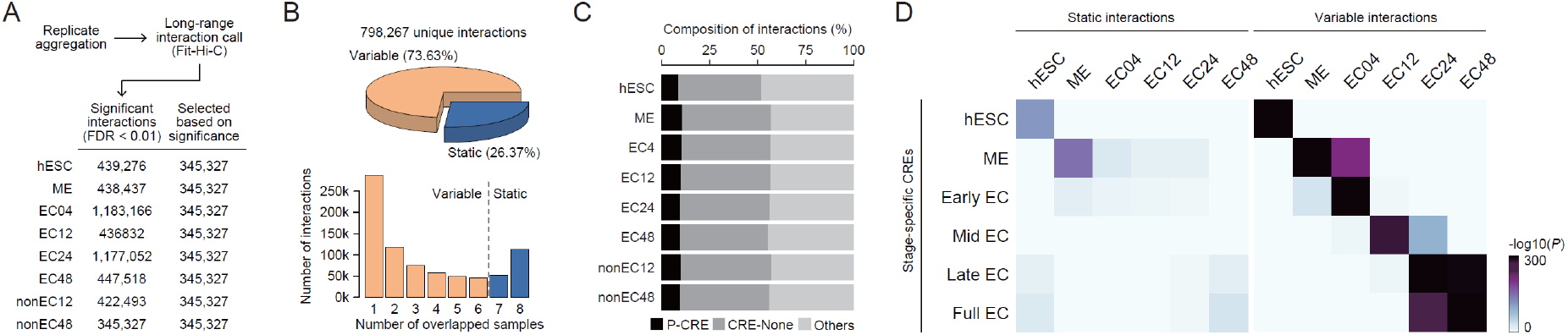
Dynamic long-range chromatin interactions during EC-differentiation. **(A)** The number of identified significant interactions during EC-differentiation. To address the undersampling problem, we selected the same number of top-ranked long-range chromatin interactions for each stage. **(B)** Shown are the fraction of variable and static long-range interactions (top) and the number of unique interactions in terms of the shared samples (bottom). (**C**) Barplots showing the fraction of promoter-CRE (p-CRE), CRE-none, and remaining interactions. **(D)** Heatmaps showing for hypergeometric p-values of the enrichment of EC-differentiation specific CREs at static (left) and variable (right) long-range chromatin interactions.

We further categorized these long-range interactions based on the presence of CREs at either or both anchors of the interacting regions (**Fig. 2C**). Consequently, 8.37% of long-range interactions were classified as promoter-CRE interactions, while 44.11% were categorized as CRE-none interactions. Next, we assessed whether variable promoter-CRE or CRE-none interactions were coordinated with stage-specific CRE activities. The hypergeometric p-value test demonstrated significant overlap between the stage-specific CREs and variable long-range interactions (**Fig. 2D**). Our findings strongly support the idea that differentiation-specific long-range chromatin interactions, primarily linked by CREs, play a critical role in EC specification.

### A development of GEM-Finder to link GWAS variants to CREs connecting stage-specific genes

Many recent studies have demonstrated that stage-specific CREs are crucial for characterizing non-coding genetic variants. Mapping GWAS variants to cell-type/differentiation-specific CREs has significantly enhanced our understanding of complex human diseases/traits by identifying disease-relevant cell types and target genes of non-coding variants [30][31]. However, a substantial portion of GWAS variants remains unexplained, as many are located outside of or in non-stage-specific CREs. Additionally, not all cell-type or stage-specific CREs are linked to promoter regions, and not all common or stage-shared CREs are unimportant to understand stage-specific regulatory systems, which limits the functional interpretation of CREs harboring GWAS variants.

To address these limitations, we hypothesized that analyzing candidate CREs connected to stage-specific genes through long-range chromatin interactions would provide greater insights into GWAS variants. To test our hypothesis, we proposed a new analytical framework called <u>G</u>enomic <u>E</u>lement <u>M</u>apping for <u>Fin</u>e Discov<u>er</u>y of Promoter-Linked Variants (GEM-Finder) (**Fig. 3A**). We applied GEM-Finder to investigate associations between complex human diseases/traits and EC differentiation, as many complex human diseases are believed to be linked to the cellular dedifferentiation process. We collected GWAS SNPs from 541 human diseases/traits and tested their associations with EC differentiation stages. The enrichment analysis assessed the relationship of selected GWAS variants with either DEG-linked CREs or stage-specific CREs. The stage-specific DEG-linked CREs were derived from promoter-CRE long-range chromatin interactions of DEGs. Next, GEM-Finder extracted DEG-linked CREs, derived from promoter-CRE long-range chromatin interactions of DEGs. For each stage, we identified approximately 8,500–36,000 DEG-linked CREs (8,490 for early, 33,092 for mid, 36,280 for late, and 19,622 for full differentiation stages). Notably, using GEM-Finder, we identified 53 disease/trait associations in total during the EC differentiation stage through a permutation test, which is 7.6 times better than those identified by stage-specific CREs (7 diseases/traits were captured) (**Fig. 3B**).

**Fig. 3.**
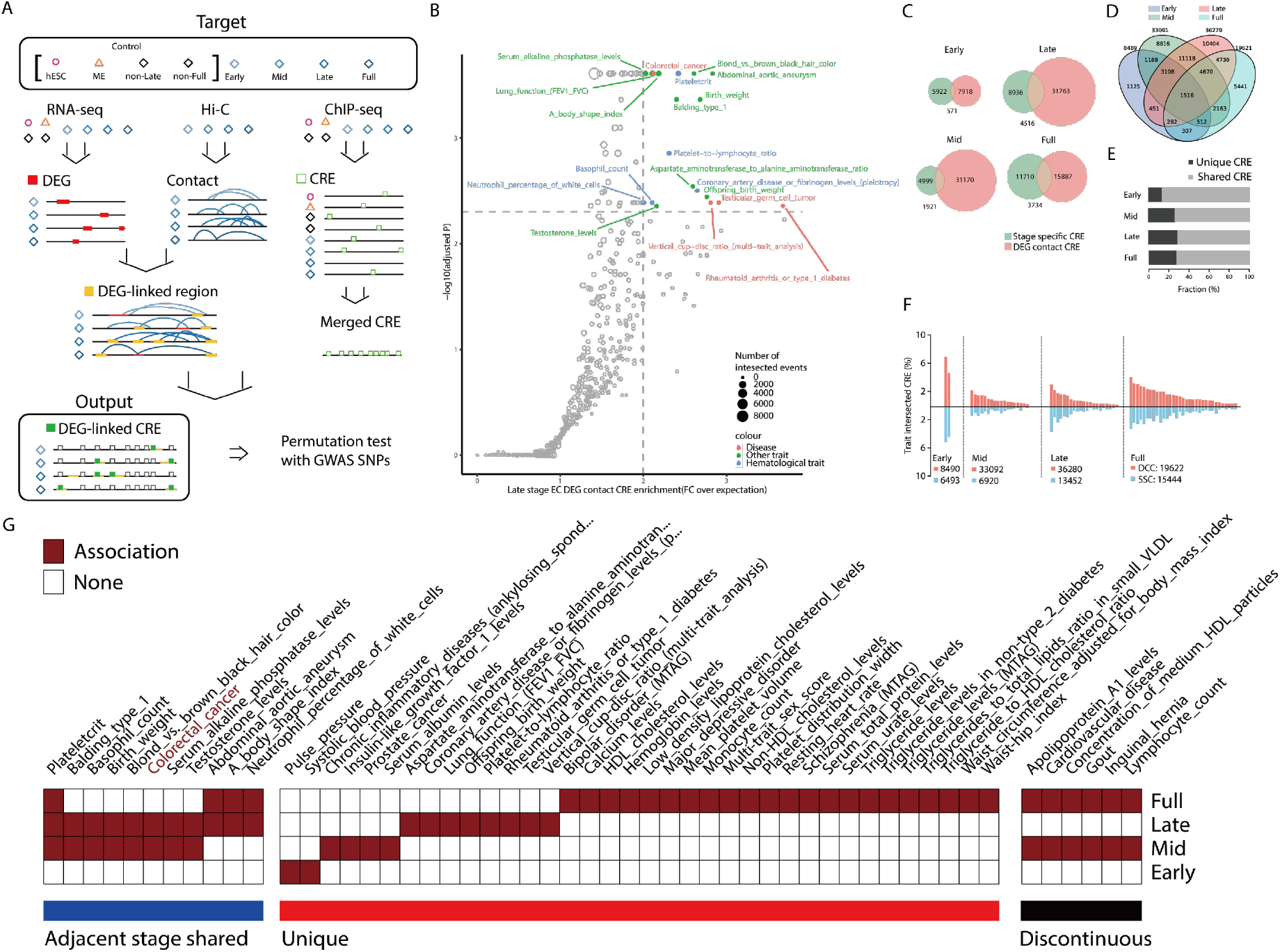
Development of GEM-Finder and its application to EC-differentiation. **(A)** Overview of the GEM-Finder workflow. **(B)** Scatter plot presents permutation results with late stage DEG-linked CRE and 541 diseases/traits. Colored are significantly associated traits/diseases with Late EC. **(C)** Venn-diagrams show overlapping CREs between H3K27ac activity-based stage-specific CREs and DEG-linked CREs. **(D)** 4-circle Venn-diagram shows overlapping DEG-linked CREs among four differentiation stages. **(E)** Barplots highlight fractions of uniquely appeared DEG-linked CRE in each stage, and shared ones. **(F)** Bi-directional plots compare ratio of CREs harboring GWAS variants between stage specific CREs and DEG-linked CREs. **(G)** Heatmap represents associated stages of each disease/trait. Clustered by (1) shared by adjacent stages, (2) uniquely associated at specific stage, and (3) discontinuously associated at multi-stages.

To elucidate the enhanced performance of GEM-Finder, we examined how many stage-specific CREs are included in DEG-linked CREs. We found that only 10–30% were included, indicating that many stage-specific CREs identified by ChIP-seq data do not directly connect to matched stages’ specific promoter regions (**Fig. 3C**). In addition, DEG-linked CREs revealed that only a small proportion of them were specifically associated with one differentiation stage (**Fig. 3D**). Approximately 10–30% of DEG-linked CREs are specifically related with one stage, and 60–80% of DEG-linked CREs were shared by more than one stages (**Fig. 3E**). Our results suggest that the strict standard for defining stage specific CREs could distinguish each stage well, but they couldn’t effectively reflect stage-specific gene regulatory programs due to loss of lots of information. The involvement of shared CREs in stage-specific DEGs may reflect complex gene regulatory programs.

Next, we examined the fraction of CREs co-localized with LD-expanded GWAS variants by GEM-Finder. We found that DEG-linked CREs displayed similar trends of GWAS variant enrichment across diseases/traits like stage-specific CREs, but a higher proportion of DEG-linked CREs were co-localized with GWAS variants (**Fig. 3F**). This result suggests that DEG-linked CREs may help mitigate false negatives from stage-specific analyses by leveraging long-range chromatin interactions and including additional DEG-linked CREs harboring GWAS variants.

When we focused on 53 significantly EC-associated diseases/traits, there were some notable findings (**Fig. 3G**). Take it for granted; many hematological traits, which are child traits of hematological measurement in the GWAS catalog, make significant associations with the EC, suggesting a significant role for EC differentiation-specific gene regulation in blood composition. In terms of stage-specific associations, we categorized these diseases/traits into three groups: those shared by adjacent stages, those uniquely associated traits, and those shared by discontinuous stages. Interestingly, 68% of EC-associated diseases/traits were uniquely linked to a specific differentiation stage, while the inclusion of adjacent stages accounted for 89% of the associations. In this organized result, we could get much information. For instance, only two traits are associated with the early stage, which are all blood pressure traits (pulse pressure and systolic blood pressure), and other 8 tested blood pressure-related traits are not observed in other differentiation stages. This result suggests that the early stage-specific regulatory program will be a critical factor in determining the risk of blood pressure. The full stage showed associations not only with hematological properties but also with neuropsychiatric diseases and diabetes. The result aligns with recent studies highlighting potential blood-brain barrier dysfunctions in psychotic disorders [32][33]. We also provided new biological insights by revealing a genomic link between EC differentiation and susceptibility to autoimmune disorders and colorectal cancer, evidenced by significant colocalization of DEG-linked CREs with their respective GWAS variants (**Fig. 3G**). The overrepresentation of these traits and diseases in EC’s gene regulation programs aligns with the physiological functions of the endothelium, as its dysfunction is likely to impact vascular function, immune cell trafficking, and tissue microenvironments [15-16, 19-20]. Overall, GEM-Finder provided more associations with complex human diseases/traits compared with ChIP-seq based stage-specific CRE-based methods.

## DISCUSSION

In this study, we developed GEM-Finder and applied it to multi-omics data (transcriptome, epigenome, and 3D genome) during endothelial cell (EC) differentiation, identifying stage-specific associations with complex human diseases/traits. GEM-Finder utilized long-range chromatin interactions and collected CREs connected to the genes with stage-specificity, revealing a novel insight into the genetic links between differentiation-dependent *cis*-gene regulatory networks and specific pathological dysfunctions. Unlike traditional methods that focus solely on differentiation-specific CREs, our approach enhances the ability to link a broader range of diseases/traits to GWAS variants. The significant increase in the number of associated complex diseases and traits highlights that both cell-type/differentiation-specific CREs and shared CREs are critical for the precise interpretation of GWAS variants. Although we are aware that integrating multi-omics resources adds complexity to applying GEM-Finder, the functional interpretation of GWAS SNPs in non-coding regions remains a challenge. In particular, while many related multi-omics resources are available, prioritizing disease-relevant cell types or differentiation stages remains an unsolved question. Given the rapid accumulation of vast amounts of differentiation-related multi-omics resources, we believe that GEM-Finder will provide a valuable approach for gaining insights into this challenge.

We also discovered numerous EC stage-specific associations for each disease, extending the previous understanding of cell-type associations to the level of cellular differentiation. Particularly, variants associated with colorectal cancer were exclusively enriched in later-stage specific DEG-linked CREs, suggesting that tumor-associated endothelial cell (TEC) formation is closely linked to the “re-activation” of differentiated ECs, likely triggered by the hypoxic and high-VEGF-A tumor microenvironment.

Note that this framework is in an initial step, so lots of strategies could improve GEM-Finder’s performance. Without delving into parameter and threshold optimization, it’s clear that developing a novel prioritization method that incorporates both stage-specific CREs and DEG-linked CREs could further enhance the functional characterization of GWAS variants. Although there are few overlapped CREs between stage-specific CREs and DEG-linked CREs **(Fig. 3C)**, the co-localization fractions of GWAS variants are quite similar **(Fig. 3F)**, suggesting that both groups contain differentiation-stage specific information. Given the complexity of the gene regulatory network, it is impossible to represent the entire cis-regulatory network for a given stage using only differentiation stage-specific CREs. Therefore, combining both CRE groups can help address false positive and false negative associations. The last but not least comment is that GEM-Finder is not limited to a specific differentiation model; rather, it can be applied to any cell type to prioritize disease-relevant cell types or differentiation stages.

To conclude, GEM-Finder clearly demonstrated the utility of long-range chromatin interactions for functional specification of non-coding variants associated with diverse human diseases/traits.

## Data Availability

All raw sequencing data used in this study have been obtained from the GEO repository (GSE201709).

## Acknowledgments

We thank the members of the Jung laboratory who provided support and suggestions throughout the course of this work.

## Author contributions

G.P., A.J.L. and I.J. designed and conceived the study. G.P. and A.J.L. analyzed and interpreted the results. G.P. and A.J.L. prepared the manuscript with assistance from I.J. All authors read and approved the final manuscript.

## Data and materials availability

Hi-C data is in GEO (GSE281670). RNA-seq and ChIP-seq data were obtained from an unpublished study via personal communication. Custom code supporting this work has been deposited in the GitHub repository (https://github.com/Delicate5/GEM-Finder). Any additional information required to reanalyze the data reported in this study is available from the lead contact upon request.

